# SpO_2_/FiO_2_ Ratio as a Better Metric for Assessment of RBC Transfusion Effectiveness in Non-traumatic Critically Ill Patients with Physiologic Derangements

**DOI:** 10.1101/2024.10.03.24313829

**Authors:** Tilendra Choudhary, Geoffrey Smith, John D. Roback, Ravi M. Patel, Cassandra D. Josephson, Rishikesan Kamaleswaran

**Affiliations:** Duke University School of Medicine, Durham NC; Emory University School of Medicine, Atlanta GA; Department of Oncology, The Johns Hopkins University School of Medicine, Baltimore, Maryland, USA; Cancer and Blood Disorders Institute, Johns Hopkins All Children’s Hospital, St. Petersburg, Florida, USA

**Author notes:** **Corresponding author:** Tilendra Choudhary, 2301 Erwin Rd, Durham, NC 27710. **E-mail addresses:** (Geoffrey Smith), (John D. Roback), (Ravi M. Patel), (Cassandra D. Josephson), (Rishikesan Kamaleswaran).

**Keywords:** red blood cell transfusion, hemoglobin, SpO_2_, SF ratio, transfusion response, critical ill patients

## Abstract

Identifying critically ill patients who are likely to improve their respiratory physiology following RBC blood transfusion is dynamic and difficult. Current decision tools are over-reliant on hemoglobin transfusion thresholds, without considering respiratory measures that may reflect physiologic effects of anemia and functional responses to RBC transfusion. Further, routine clinical measures to determine transfusion efficacy beyond hemoglobin increment are lacking to identify patients as responders or non-responders. We present a two-center retrospective cohort study aiming to determine a potential biomarker to assess the physiologic response of RBC transfusion for ICU patients. The study was performed with 13,274 eligible admissions at the first center. Another 3,757 from the second center were used for validation purpose. We introduced a comparative analysis of two respiratory measures, SpO_2_ and SpO_2_/FiO_2_ (SF) ratio, in addition to hemoglobin, to assess individual patient responses to RBC transfusion. A statistical study was performed to analyze the change in these markers before and after the transfusion interval. Based on quantitative statistical analyses, we found SF ratio to be a more effective biomarker than hemoglobin alone for revealing RBC transfusion efficacy. There existed an inverse correlation between pre-transfusion SF and transfusion efficacy. The results were consistent across both centers, revealing generalizability.

## Introduction

Critical ill patients in intensive care units (ICU) often develop anemia and coagulation that are associated with adverse outcomes, such as high risk of life-threatening scenarios, thrombosis, and coronary artery diseases. To treat the anemia and coagulation following surgery, trauma or medical conditions, patients have been transfused with red blood cells (RBCs) for decades [1,2]. Annually about 85 million RBC units are used for transfusion worldwide, and approximately 15 million are transfused every year in the United States only. In transfusion medicine, a major physical injury or massive bleeding due to an accident, surgery, or uncontrolled intraoperative hemorrhage is typically referred as *Trauma*. Massive blood transfusions (MTs) are essential for traumatic patients. In contrast, reasons for non-traumatic blood transfusions include healthy blood cell deficiency, anemia, coagulation, and other health disorders (e.g., hemophilia, thrombocytopenia, kidney or liver disease, severe infection, and sickle cell disease). An assessment of transfusion response and transfusion necessity in non-traumatic ICU patients is more difficult than in traumatic patients. The hallmark trigger for initiating transfusions has been the criterion of hemoglobin (Hb) ≤7 g/dL [1–3]. However, a significant percentage of transfusions performed are often identified as inappropriate [4,5], thus contributing to adverse events such as acute immune hemolytic reactions and bloodborne infections [6–8]. There has been much interest in the discovery of alternative biomarkers that may indicate a quantitative utility of RBC transfusions. One potential non-invasive derived measure is the SpO_2_/FiO_2_ (SF) ratio, which incorporates blood oxygen content along with oxygen concentration in gas exchange. This equation couples the intensity of respiratory support, the patient is receiving – FiO_2_, with the degree of hypoxemia – SpO_2_, the patient is experiencing. FiO_2_ is a measure of inhaled atmospheric/supplemental oxygen content and can be titrated according to the SpO_2_ measure. When oxygen supply and its consumption are incongruent, cells are damaged and subsequently die [9]. For instance, in chronic obstructive pulmonary disease (COPD), supplemental oxygen is recommended to start when the SpO_2_ falls below 88% [9]. Clinically, SF<144 is marked severe, 144-235 moderate, 235-315 mild, and >315 normal [10]. Based on the potential for the SF ratio to provide a functional measure of oxygen requirements and delivery, the SF was hypothesized to be a more accurate biomarker for RBC transfusion response. This study endeavored to investigate the effect of RBC transfusion on SF ratio in a cohort of critically ill patients to determine the physiologic response to RBC transfusion. The study may help in devising decision-making tools, data benchmarking, and future research to optimize treatment strategies for RBC transfusion of ICU patients.

## Materials and Methods

This study was reviewed and approved by the Emory University IRB (No. STUDY00000302) as non-human subjects, and waiver of informed consent was obtained. All procedures were followed in accordance with the ethical standards of the responsible committee on human experimentation (institutional/regional) and with the Helsinki Declaration of 1975. Continuous physiological data were archived using the BedMaster (Excel Medical, Jupiter, FL) software. SpO_2_ was collected at 1-hour sampling interval and used to derive the SF ratio continuously from admission through to discharge for patients. Hemoglobin and FiO_2_ values were derived from the electronic medical record for enrolled patients. The retrospective data was lastly accessed on Mar. 30^th^, 2024, for this research. The authors had also access to information that could identify individual participants during or after data collection.

In this article, we present a multi-center retrospective cohort study conducted at two high volume academic hospitals located in the southeastern U.S.A. Adult non-traumatic ICU patients (≥18 years) admitted to either of these two metropolitan hospitals, Emory University Hospital and Grady Memorial Hospital (Atlanta, GA) anytime in 2016-2020, who received RBC transfusion, were eligible for enrollment in the study. We adopted a multi-center derivation and validation study design by first establishing relation of SF and transfusion response using data from Emory University Hospital, and then validating this relation against data collected from Grady Memorial Hospital. Patients were excluded for the following reasons: a) if they had trauma and high bleeding injury who received massive transfusions, b) if they did not have continuous monitoring data archived during encounter, c) were discharged or died after ICU admission within 24 hours, due to limited duration of monitoring data available; and, d) who did not have continuous physiological data up to 24 hours before and after the transfusion. We also excluded successive RBC transfusion episodes occurred within next 24 hours of an RBC transfusion event to avoid data overlapping.

We divided the timing of RBC transfusion into two windows, the period before the initiation of RBC transfusion and the period after its completion. Specifically, 24-hour data sequences were considered for the period before transfusion, and three subsequent post-RBC transfusion periods (T1, T2, T3; 9-hour duration each) were evaluated for the post-transfusion period, typically following a 4-hour period of RBC infusion. We applied median aggregation to the data of our cohort, across the pre-transfusion window and post-transfusion time-windows. An overview of our method is shown in Figure 1. The reason for the selection of the three periods was to identify short-term vs. sustained improvements in respiratory physiology. Responses to RBC transfusions were characterized by SF differences (ΔSF) that were calculated between post- and pre-transfusion periods, considering four different ranges of baseline SF measurements (1-100, 101-200, 201-300, >300). An increase in the SF ratio indicated improvement in respiratory physiology (either from an SpO_2_ improvement or an FiO_2_ decrease). We further employed a polynomial curve fitting scheme to evaluate the relation between baseline SF and ΔSF, demonstrating which patient groups were more benefitted from transfusion. The following third-order polynomial function was used to model this relation in a least square sense.

**Figure 1:**
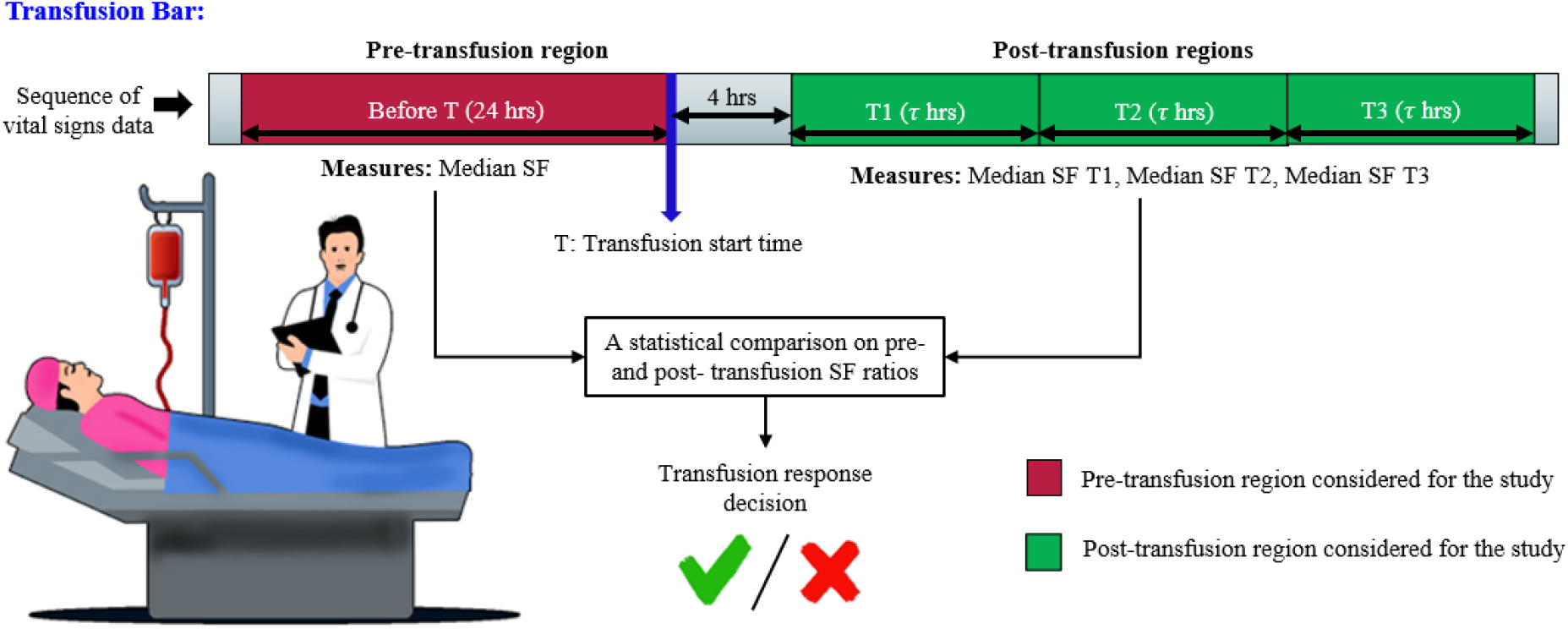
Overview of the study design. One pre-transfusion period of 24 hours and three post-transfusion periods of 9 hours each were used to analyze short-term vs. sustained physiological improvements.

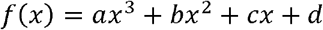

where *a, b, c, d* denote the modelling parameters. The association between ΔSF and difference in SpO_2_ (ΔSpO_2_) was also evaluated using the Bland Altman analysis.

To evaluate the statistical significance (*p*-value) among four baseline SF groups, Chi-squared and One-way ANOVA tests were used for categorical and numeric features, respectively. Whereas Pearson coefficient (r) and R-squared (R^2^) values were used for evaluating correlation.

## Results

We retrospectively evaluated a total of 82,767 patient admissions from 64,962 unique patients who were admitted to Emory ICUs from 2016-2020, of which 15,898 (∼20%) received at least one RBC transfusion with no massive transfusions. Among these, 14,765 had continuous physiologic bedside monitoring data within the temporal period close to the index RBC transfusion event. After discarding patients with missing physiological data, 13,274 were selected for the analysis and a total of 18,716 transfusion episodes were noted for which the baseline SF is not missing. Our cohort consists of transfused patients across a wide range of demographic features, such as age (mean: 61.4 ± 16 years), sex (male: 48.1%), and race (Caucasian: 46.6%, African American: 45%). Similarly, from the Grady Hospital, a total of 6,375 transfusion episodes from 3,757 patient encounters were included in the study. Clinical characteristics and demographics of transfusion patients from both the centers are listed in Tables 1 and 2 for their pre-transfusion observational data.

**Table 1:**
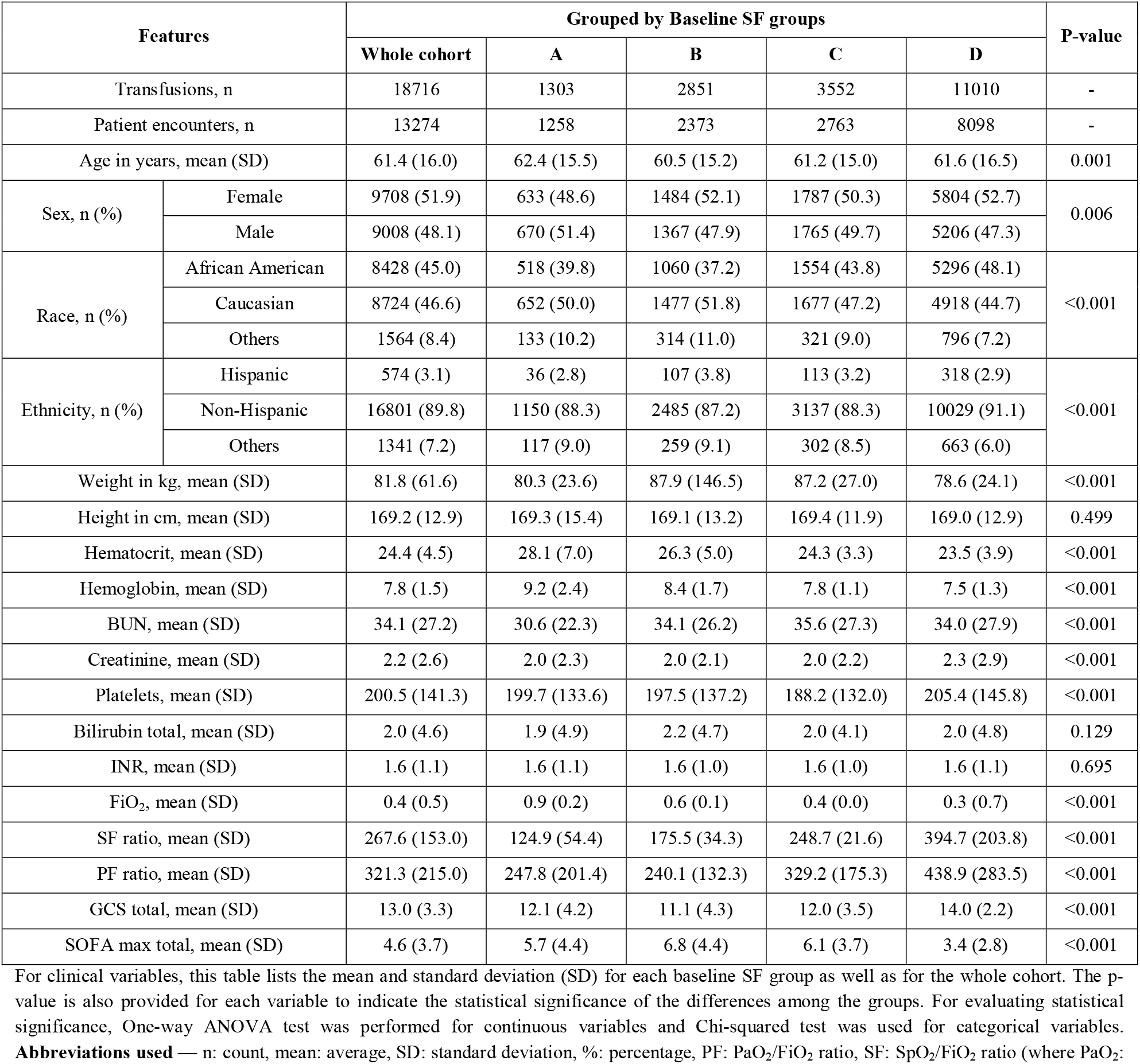
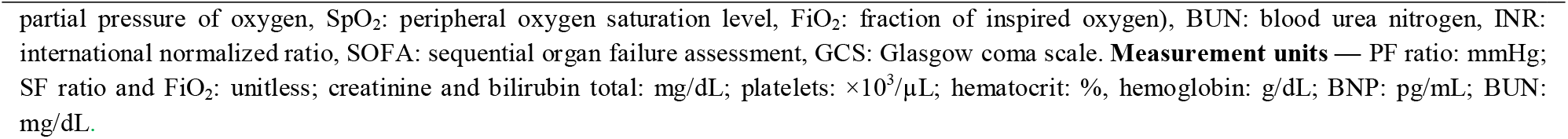
Clinical characteristics and demographic variables of the derivation dataset collected from Emory admitted RBC transfused patients based on their pre-transfusion aggregated features in four baseline SF groups – A: 1-100, B: 101-200, C: 201-300, D: >300.

| Features |  | Grouped by Baseline SF groups |  |  |  |  | P-value |
| --- | --- | --- | --- | --- | --- | --- | --- |
|  |  | Whole cohort | A | B | C | D |  |
| Transfusions, n |  | 18716 | 1303 | 2851 | 3552 | 11010 | - |
| Patient encounters, n |  | 13274 | 1258 | 2373 | 2763 | 8098 | - |
| Age in years, mean (SD) |  | 61.4 (16.0) | 62.4 (15.5) | 60.5 (15.2) | 61.2 (15.0) | 61.6 (16.5) | 0.001 |
| Sex, n (%) | Female | 9708 (51.9) | 633 (48.6) | 1484 (52.1) | 1787 (50.3) | 5804 (52.7) | 0.006 |
|  | Male | 9008 (48.1) | 670 (51.4) | 1367 (47.9) | 1765 (49.7) | 5206 (47.3) |  |
| Race, n (%) | African American | 8428 (45.0) | 518 (39.8) | 1060 (37.2) | 1554 (43.8) | 5296 (48.1) | <0.001 |
|  | Caucasian | 8724 (46.6) | 652 (50.0) | 1477 (51.8) | 1677 (47.2) | 4918 (44.7) |  |
|  | Others | 1564 (8.4) | 133 (10.2) | 314 (11.0) | 321 (9.0) | 796 (7.2) |  |
| Ethnicity, n (%) | Hispanic | 574 (3.1) | 36 (2.8) | 107 (3.8) | 113 (3.2) | 318 (2.9) | <0.001 |
|  | Non-Hispanic | 16801 (89.8) | 1150 (88.3) | 2485 (87.2) | 3137 (88.3) | 10029 (91.1) |  |
|  | Others | 1341 (7.2) | 117 (9.0) | 259 (9.1) | 302 (8.5) | 663 (6.0) |  |
| Weight in kg, mean (SD) |  | 81.8 (61.6) | 80.3 (23.6) | 87.9 (146.5) | 87.2 (27.0) | 78.6 (24.1) | <0.001 |
| Height in cm, mean (SD) |  | 169.2 (12.9) | 169.3 (15.4) | 169.1 (13.2) | 169.4 (11.9) | 169.0 (12.9) | 0.499 |
| Hematocrit, mean (SD) |  | 24.4 (4.5) | 28.1 (7.0) | 26.3 (5.0) | 24.3 (3.3) | 23.5 (3.9) | <0.001 |
| Hemoglobin, mean (SD) |  | 7.8 (1.5) | 9.2 (2.4) | 8.4 (1.7) | 7.8 (1.1) | 7.5 (1.3) | <0.001 |
| BUN, mean (SD) |  | 34.1 (27.2) | 30.6 (22.3) | 34.1 (26.2) | 35.6 (27.3) | 34.0 (27.9) | <0.001 |
| Creatinine, mean (SD) |  | 2.2 (2.6) | 2.0 (2.3) | 2.0 (2.1) | 2.0 (2.2) | 2.3 (2.9) | <0.001 |
| Platelets, mean (SD) |  | 200.5 (141.3) | 199.7 (133.6) | 197.5 (137.2) | 188.2 (132.0) | 205.4 (145.8) | <0.001 |
| Bilirubin total, mean (SD) |  | 2.0 (4.6) | 1.9 (4.9) | 2.2 (4.7) | 2.0 (4.1) | 2.0 (4.8) | 0.129 |
| INR, mean (SD) |  | 1.6 (1.1) | 1.6 (1.1) | 1.6 (1.0) | 1.6 (1.0) | 1.6 (1.1) | 0.695 |
| FiO<sub>2</sub>, mean (SD) |  | 0.4 (0.5) | 0.9 (0.2) | 0.6 (0.1) | 0.4 (0.0) | 0.3 (0.7) | <0.001 |
| SF ratio, mean (SD) |  | 267.6 (153.0) | 124.9 (54.4) | 175.5 (34.3) | 248.7 (21.6) | 394.7 (203.8) | <0.001 |
| PF ratio, mean (SD) |  | 321.3 (215.0) | 247.8 (201.4) | 240.1 (132.3) | 329.2 (175.3) | 438.9 (283.5) | <0.001 |
| GCS total, mean (SD) |  | 13.0 (3.3) | 12.1 (4.2) | 11.1 (4.3) | 12.0 (3.5) | 14.0 (2.2) | <0.001 |
| SOFA max total, mean (SD) |  | 4.6 (3.7) | 5.7 (4.4) | 6.8 (4.4) | 6.1 (3.7) | 3.4 (2.8) | <0.001 |

For clinical variables, this table lists the mean and standard deviation (SD) for each baseline SF group as well as for the whole cohort. The p-value is also provided for each variable to indicate the statistical significance of the differences among the groups. For evaluating statistical significance, One-way ANOVA test was performed for continuous variables and Chi-squared test was used for categorical variables. **Abbreviations used** — n: count, mean: average, SD: standard deviation, %: percentage, PF: PaO<sub>2</sub>/FiO<sub>2</sub> ratio, SF: SpO<sub>2</sub>/FiO<sub>2</sub> ratio (where PaO<sub>2</sub>:
partial pressure of oxygen, SpO<sub>2</sub>: peripheral oxygen saturation level, FiO<sub>2</sub>: fraction of inspired oxygen), BUN: blood urea nitrogen, INR: international normalized ratio, SOFA: sequential organ failure assessment, GCS: Glasgow coma scale. **Measurement units** — PF ratio: mmHg; SF ratio and FiO<sub>2</sub>: unitless; creatinine and bilirubin total: mg/dL; platelets: $\times 10^3/\mu\text{L}$ ; hematocrit: %, hemoglobin: g/dL; BNP: pg/mL; BUN: mg/dL.

**Table 2:**
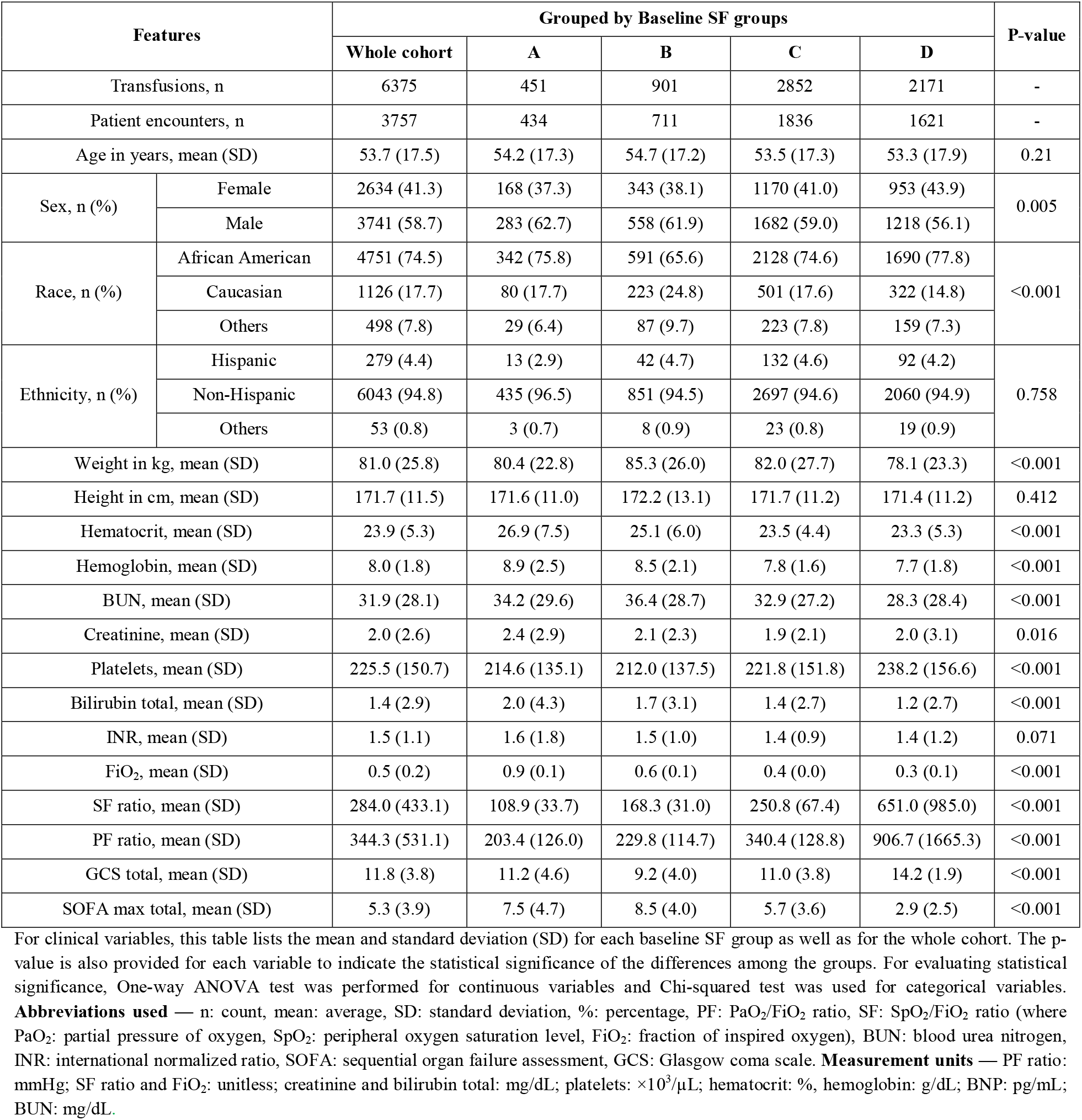
Clinical characteristics and demographic variables of the validation dataset collected from Grady admitted RBC transfused patients based on their pre-transfusion aggregated features in four baseline SF groups – A: 1-100, B: 101-200, C: 201-300, D: >300.

| Features |  | Grouped by Baseline SF groups |  |  |  |  | P-value |
| --- | --- | --- | --- | --- | --- | --- | --- |
|  |  | Whole cohort | A | B | C | D |  |
| Transfusions, n |  | 6375 | 451 | 901 | 2852 | 2171 | - |
| Patient encounters, n |  | 3757 | 434 | 711 | 1836 | 1621 | - |
| Age in years, mean (SD) |  | 53.7 (17.5) | 54.2 (17.3) | 54.7 (17.2) | 53.5 (17.3) | 53.3 (17.9) | 0.21 |
| Sex, n (%) | Female | 2634 (41.3) | 168 (37.3) | 343 (38.1) | 1170 (41.0) | 953 (43.9) | 0.005 |
|  | Male | 3741 (58.7) | 283 (62.7) | 558 (61.9) | 1682 (59.0) | 1218 (56.1) |  |
| Race, n (%) | African American | 4751 (74.5) | 342 (75.8) | 591 (65.6) | 2128 (74.6) | 1690 (77.8) | <0.001 |
|  | Caucasian | 1126 (17.7) | 80 (17.7) | 223 (24.8) | 501 (17.6) | 322 (14.8) |  |
|  | Others | 498 (7.8) | 29 (6.4) | 87 (9.7) | 223 (7.8) | 159 (7.3) |  |
| Ethnicity, n (%) | Hispanic | 279 (4.4) | 13 (2.9) | 42 (4.7) | 132 (4.6) | 92 (4.2) | 0.758 |
|  | Non-Hispanic | 6043 (94.8) | 435 (96.5) | 851 (94.5) | 2697 (94.6) | 2060 (94.9) |  |
|  | Others | 53 (0.8) | 3 (0.7) | 8 (0.9) | 23 (0.8) | 19 (0.9) |  |
| Weight in kg, mean (SD) |  | 81.0 (25.8) | 80.4 (22.8) | 85.3 (26.0) | 82.0 (27.7) | 78.1 (23.3) | <0.001 |
| Height in cm, mean (SD) |  | 171.7 (11.5) | 171.6 (11.0) | 172.2 (13.1) | 171.7 (11.2) | 171.4 (11.2) | 0.412 |
| Hematocrit, mean (SD) |  | 23.9 (5.3) | 26.9 (7.5) | 25.1 (6.0) | 23.5 (4.4) | 23.3 (5.3) | <0.001 |
| Hemoglobin, mean (SD) |  | 8.0 (1.8) | 8.9 (2.5) | 8.5 (2.1) | 7.8 (1.6) | 7.7 (1.8) | <0.001 |
| BUN, mean (SD) |  | 31.9 (28.1) | 34.2 (29.6) | 36.4 (28.7) | 32.9 (27.2) | 28.3 (28.4) | <0.001 |
| Creatinine, mean (SD) |  | 2.0 (2.6) | 2.4 (2.9) | 2.1 (2.3) | 1.9 (2.1) | 2.0 (3.1) | 0.016 |
| Platelets, mean (SD) |  | 225.5 (150.7) | 214.6 (135.1) | 212.0 (137.5) | 221.8 (151.8) | 238.2 (156.6) | <0.001 |
| Bilirubin total, mean (SD) |  | 1.4 (2.9) | 2.0 (4.3) | 1.7 (3.1) | 1.4 (2.7) | 1.2 (2.7) | <0.001 |
| INR, mean (SD) |  | 1.5 (1.1) | 1.6 (1.8) | 1.5 (1.0) | 1.4 (0.9) | 1.4 (1.2) | 0.071 |
| FiO<sub>2</sub>, mean (SD) |  | 0.5 (0.2) | 0.9 (0.1) | 0.6 (0.1) | 0.4 (0.0) | 0.3 (0.1) | <0.001 |
| SF ratio, mean (SD) |  | 284.0 (433.1) | 108.9 (33.7) | 168.3 (31.0) | 250.8 (67.4) | 651.0 (985.0) | <0.001 |
| PF ratio, mean (SD) |  | 344.3 (531.1) | 203.4 (126.0) | 229.8 (114.7) | 340.4 (128.8) | 906.7 (1665.3) | <0.001 |
| GCS total, mean (SD) |  | 11.8 (3.8) | 11.2 (4.6) | 9.2 (4.0) | 11.0 (3.8) | 14.2 (1.9) | <0.001 |
| SOFA max total, mean (SD) |  | 5.3 (3.9) | 7.5 (4.7) | 8.5 (4.0) | 5.7 (3.6) | 2.9 (2.5) | <0.001 |

For clinical variables, this table lists the mean and standard deviation (SD) for each baseline SF group as well as for the whole cohort. The p-value is also provided for each variable to indicate the statistical significance of the differences among the groups. For evaluating statistical significance, One-way ANOVA test was performed for continuous variables and Chi-squared test was used for categorical variables. **Abbreviations used** — n: count, mean: average, SD: standard deviation, %: percentage, PF: PaO<sub>2</sub>/FiO<sub>2</sub> ratio, SF: SpO<sub>2</sub>/FiO<sub>2</sub> ratio (where PaO<sub>2</sub>: partial pressure of oxygen, SpO<sub>2</sub>: peripheral oxygen saturation level, FiO<sub>2</sub>: fraction of inspired oxygen), BUN: blood urea nitrogen, INR: international normalized ratio, SOFA: sequential organ failure assessment, GCS: Glasgow coma scale. **Measurement units** — PF ratio: mmHg; SF ratio and FiO<sub>2</sub>: unitless; creatinine and bilirubin total: mg/dL; platelets: ×10<sup>3</sup>/μL; hematocrit: %, hemoglobin: g/dL; BNP: pg/mL; BUN: mg/dL.

For the Emory data as a derivation dataset, Table 1 lists distribution of transfused patients in four groups according to their baseline SF values as A: 1-100, B: 101-200, C: 201-300, D: >300. Initially, we attempted to establish a relation between post-transfusion SF variation and baseline Hb for four different baseline SF ranges with the derivation data. For the first post-transfusion segment T1, the relation between ΔSF and baseline Hb is presented for four different baseline SF groups in Figure 2. We found that patients with the lowest baseline SF ratio (1-100), reflecting those most critically ill, consistently demonstrated an improvement in SF ratio subsequent to RBC transfusion with a mean increase of 75.9 (95% CI 71.4-80.4) after T1, 107.3 (95% CI 102.7-111.8) after T2, 123 (95% CI 118.4-127.6) after T3 (Supplemental eFigure 1). This improved effect was present across all three time periods (T1-T3), and independent of baseline Hb values (e.g. R^2^ of -0.7, -1.4 and -2.0 for T1, T2 and T3, respectively, top row of Supplemental eFigure 1). For patients with a baseline SF >300, reflecting the population with the least respiratory compromise, no significant improvement in SF ratio was observed following RBC transfusion, and in fact respiratory physiology worsened after transfusion (an average decrease of 5.9 (95% CI 5.2-6.6) after T1, 7.3 (95% CI 6.6-8.1) after T2, and 8.4 (95% CI 6.5-8.1) after T3), with mean differences actually reflecting worsening of respiratory physiology. Again, differences in SF ratios were not associated with pre-transfusion Hb values (e.g. R^2^ of -0.13, -0.14 and -0.15 for T1, T2 and T3, respectively, bottom row of Supplemental eFigure 1). Patients with more moderate baseline SF ratios had more heterogeneous responses to RBC transfusions with both positive and negative SF ratios (Supplemental eFigure 1: 2nd and 3rd rows).

**Figure 2:**
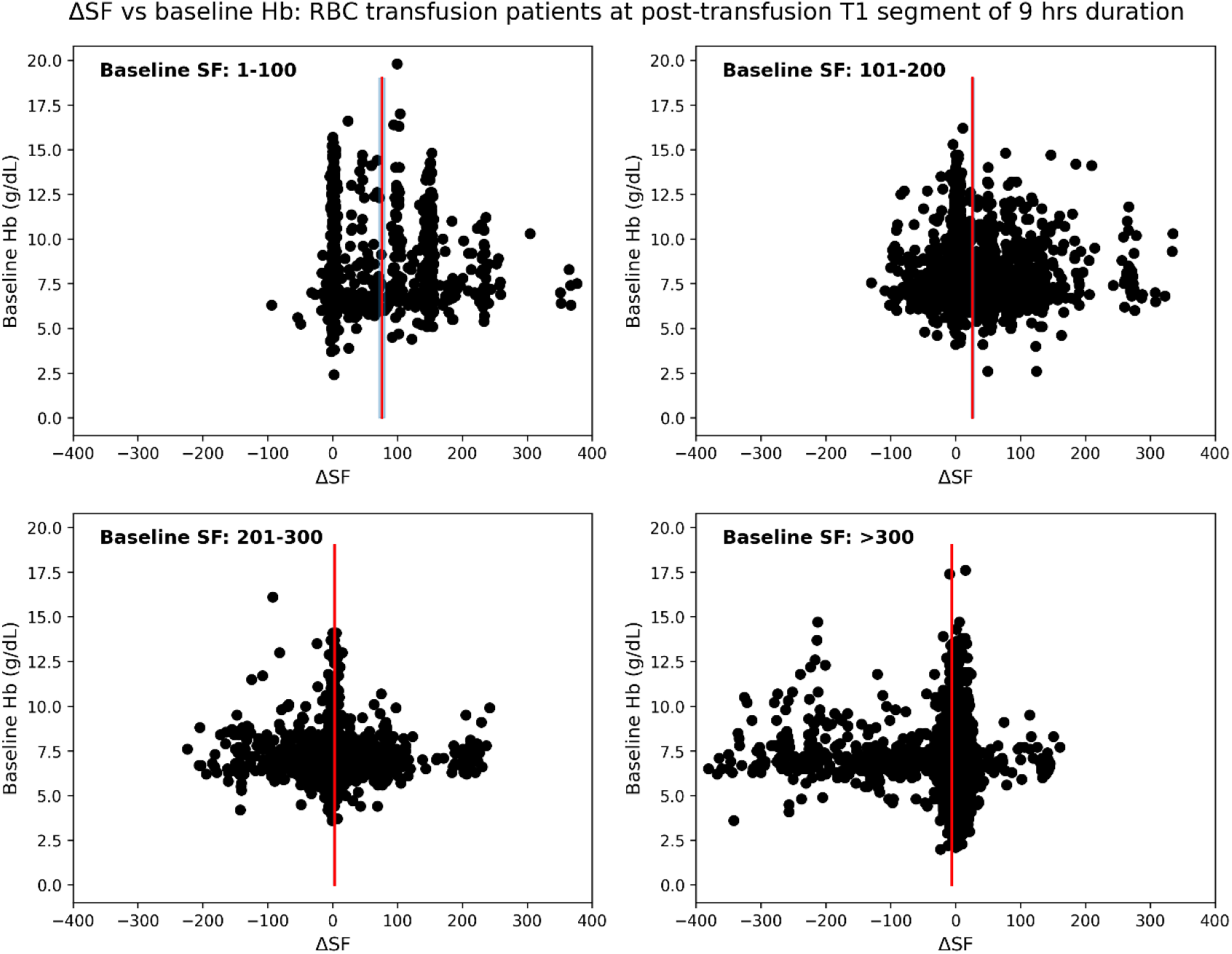
Localized study on Emory hospital patients having different ranges of baseline SF ratios. ΔSF vs baseline Hb for first post-transfusion region T1 of duration 9 hours with baseline SF range (a) 1-100, (b) 101-200, (c) 201-300, and (d) >300. Note that data-points denote the distribution of patients. Red vertical line with blueish filled region around it represents the mean difference in SF with 95% confidence interval (CI).

We also evaluated the relationship of baseline SF with RBC transfusion efficacy, expressed by ΔSF measures, using a polynomial curve fitting scheme. In this analysis, ΔSF is measured as difference in SF from pre-transfusion to first post-transfusion segment. The modelled resultant curve establishes an inverse relation of baseline SF and ΔSF, demonstrating a great transfusion benefit for patients having lower baseline SFs and the effect reduces for increasing baseline SF measure. It is shown in Figure 3. Furthermore, an association of z-score normalized ΔSF and ΔSpO_2_ was investigated using the Bland Altman and correlation analyses, which indicate a weak correlation (r=0.15, *p*<0.005) and poor agreement (95% limits of agreement LoA: [-2.6, 2.6]) for the derivation dataset. Their plots are illustrated in Figure 4.

**Figure 3:**
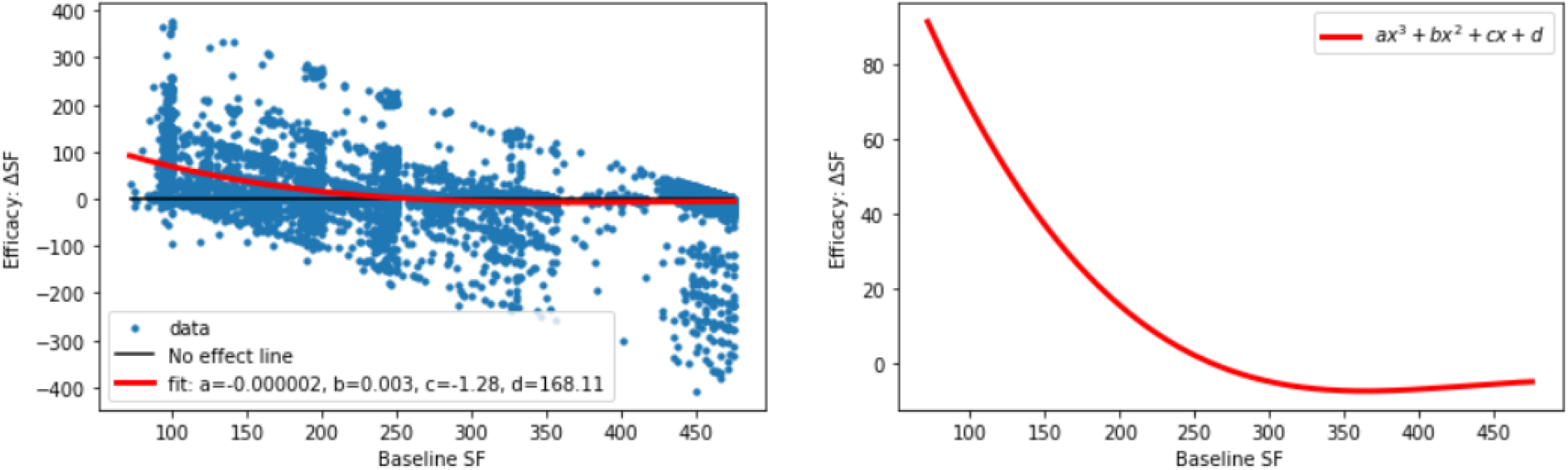
Derivation data: Relation between baseline SF (BSF) and transfusion efficacy (ΔSF). Resultant curve modelled with a polynomial curve fitting demonstrates an inverse relation of BSF and ΔSF.

**Figure 4:**
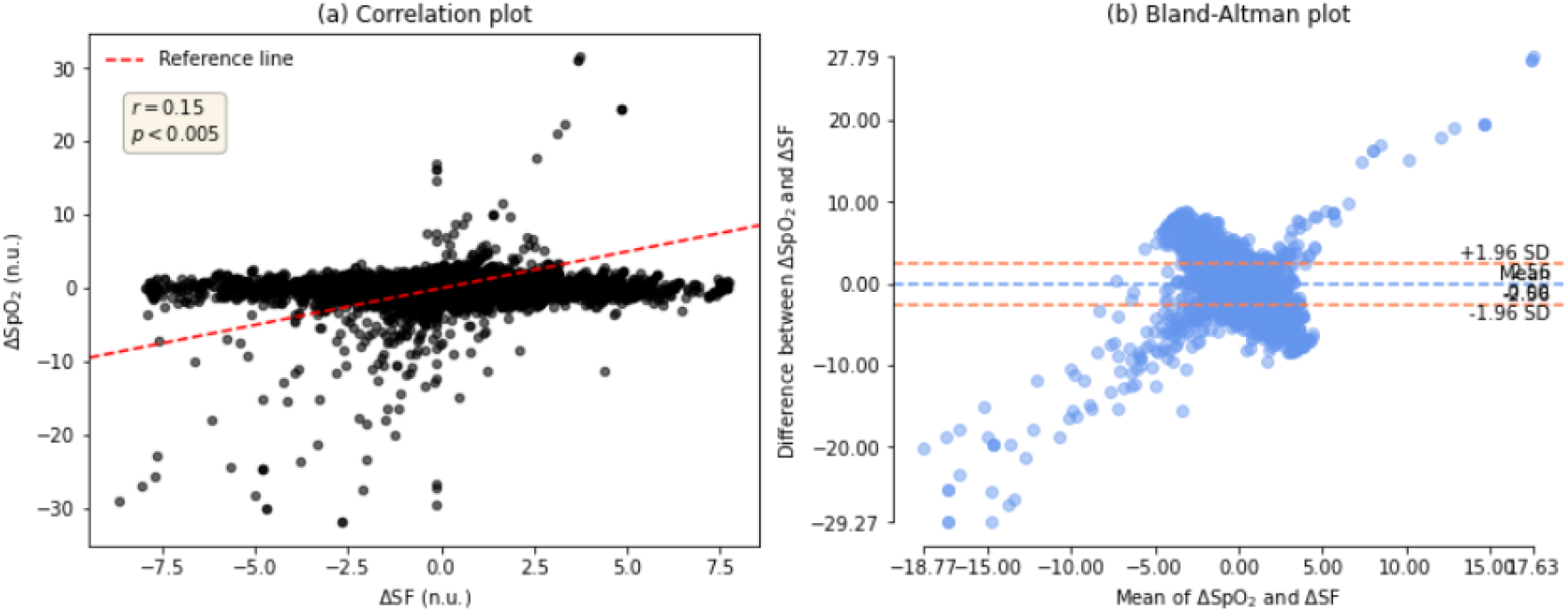
Bland Altman analysis of normalized ΔSF and ΔSpO_2_ on the derivation set. (a) correlation plot, and (b) Bland-Altman plot.

To check the consistency and reproducibility of results, we validated our method with Grady Hospital data. Similar to the derivation data, Table 2 presents the distribution of transfused patients in four baseline SF groups for the validation data. For the first post-transfusion segment T1, the relation between ΔSF and baseline Hb is shown for all four baseline SF groups in Figure 5; whereas Supplemental eFigure 2 shows a detailed picture of this relationship for T1, T2 and T3. We observed that patients with the lowest baseline SF ratio (1-100) showed an improvement in SF ratio after RBC transfusion. While patients with a baseline SF >300 demonstrated no significant improvement in post-transfusion SF ratios. These findings demonstrated a similar pattern that was obtained by the derivation data, ensuring the consistency. Additionally, the resulting modelled curve on ΔSF and baseline SF, depicted in Figure 6, showed an inverse relation of them, which was also consistent. Like for the derivation data, we also attempted to show the association of ΔSF with ΔSpO_2_ using the Bland Altman analysis in Figure 7, which also yielded poor and similar correlation (r=0.12, *p*<0.005) and agreement (LoA: [-2.61, 2.61]).

**Figure 5:**
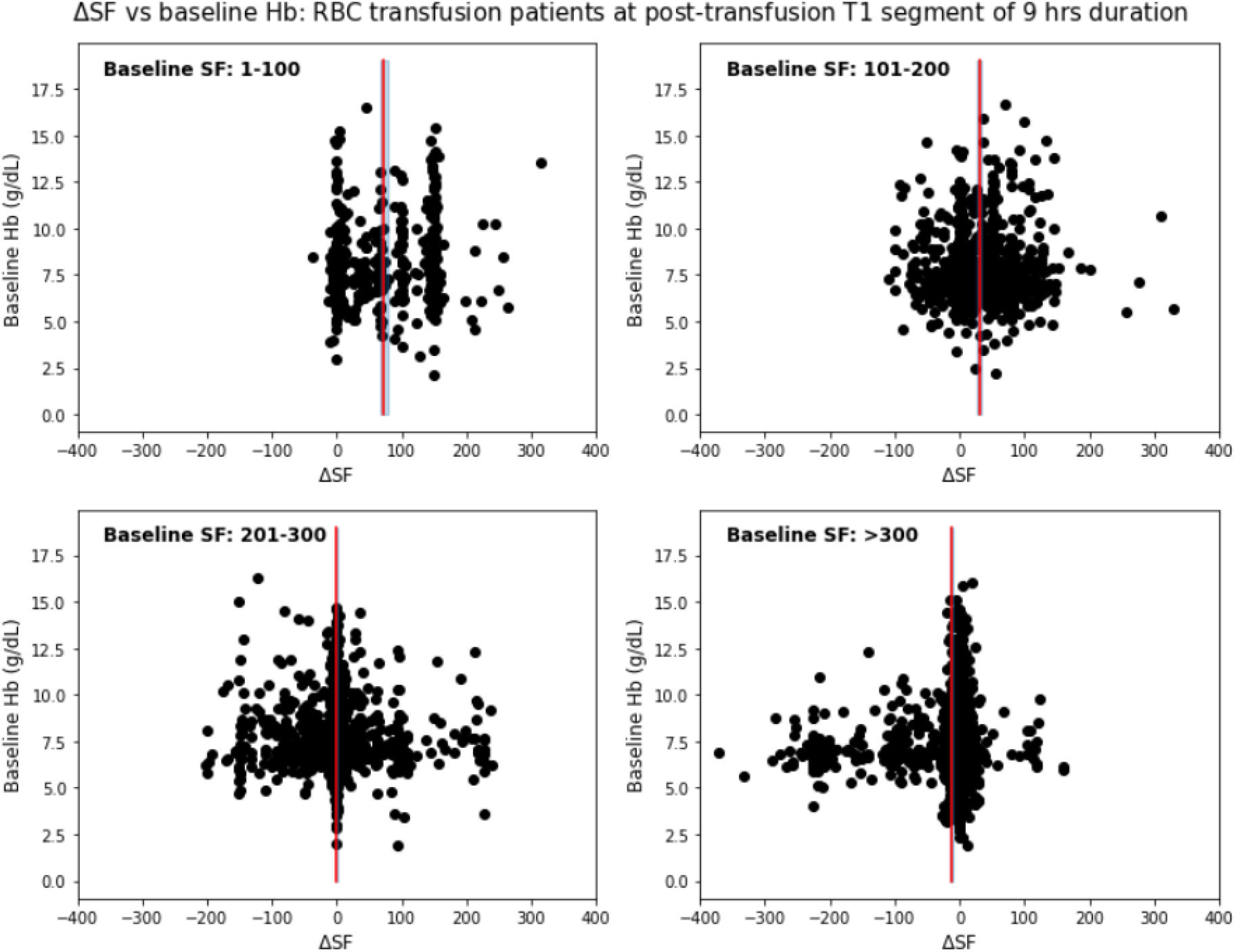
Localized study on Grady hospital patients having different ranges of baseline SF ratios. ΔSF vs baseline Hb for first post-transfusion region T1 of duration 9 hours with baseline SF range (a) 1-100, (b) 101-200, (c) 201-300, and (d) >300. Note that data-points denote the distribution of patients. Red vertical line with blueish filled region around it represents the mean difference in SF with 95% confidence interval (CI).

**Figure 6:**
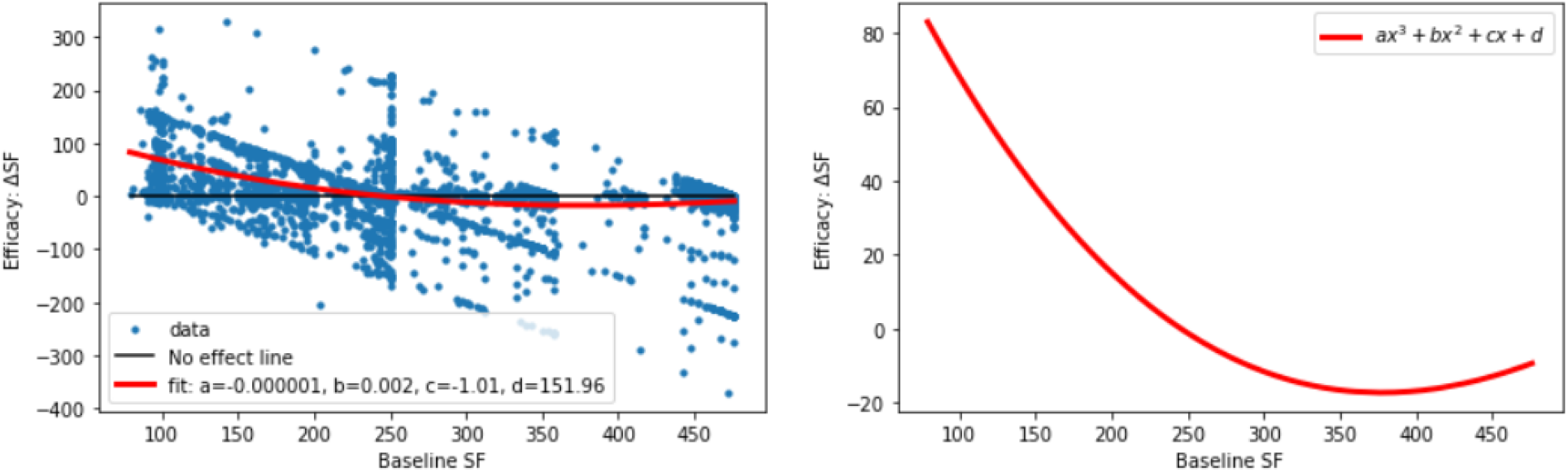
Validation data: Relation between baseline SF (BSF) and transfusion efficacy (ΔSF). Resultant curve modelled with a polynomial curve fitting demonstrates an inverse relation of BSF and ΔSF.

**Figure 7:**
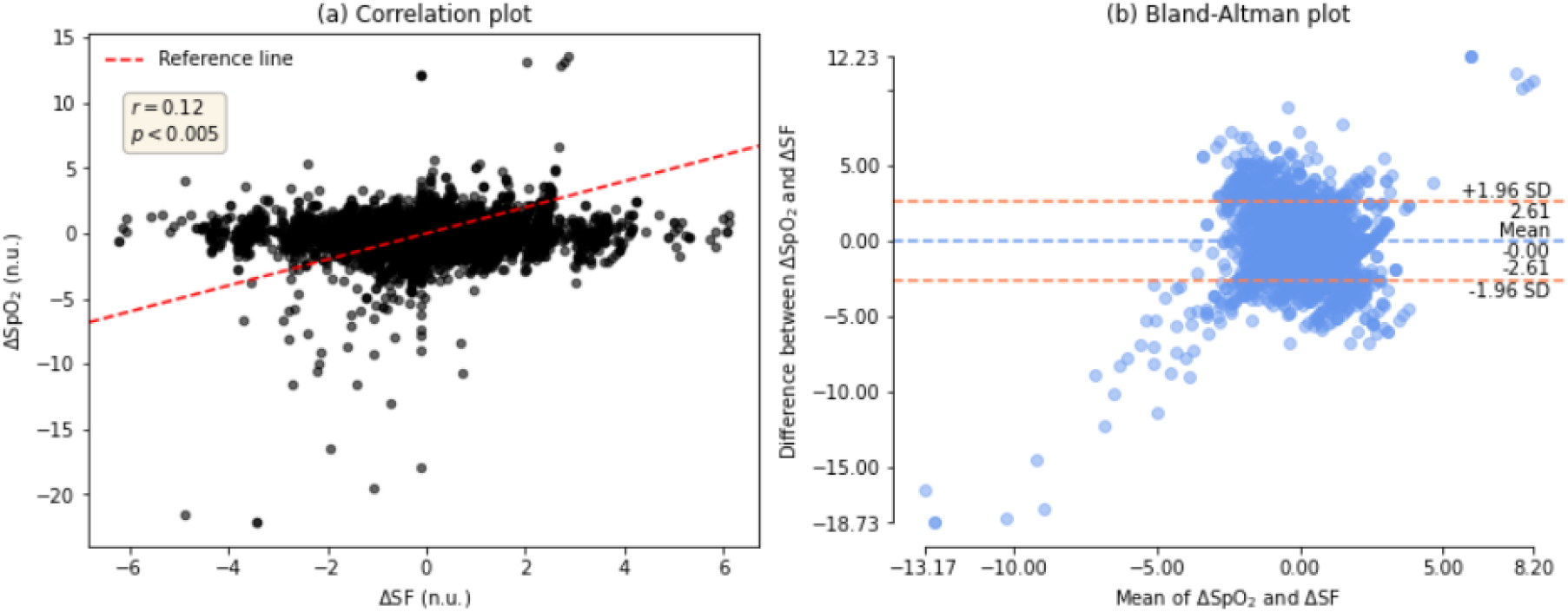
Bland Altman analysis of normalized ΔSF and ΔSpO_2_ on the validation data. (a) correlation plot, and (b) Bland-Altman plot.

## Discussion

In the present study, we demonstrated how strongly baseline SF, i.e., pre-transfusion SF ratio is associated with the RBC transfusion response, shown by ΔSF measurement, with substantial experiments on two different centers. Critically ill patients with physiologic derangements having lowest SF 1-100 benefitted more from the RBC transfusion. While respiratory physiology of population with the least respiratory compromise (baseline SF >300) worsened subsequent to RBC transfusion. Observed consistency in results across both centers, this initial study establishes SF as a potential marker to evaluate the RBC transfusion efficacy.

Figures 2, 5 and eFigures 1, 2 show almost linear and continuous decrement in post-transfusion SF change of patient-groups having incremental baseline SFs, and this effect is consistent throughout each of all three post-transfusion regions. Decrement in ΔSF is associated with the deterioration of transfusion efficacy. This effect was also shown via polynomial curve modeling approach. Also, the ΔSpO2 had poor agreement with ΔSF, highlighting that SpO2 alone may not reflect the changes in physiology informed by the more complete SF ratio.

The present study is first of its kind, specifically designed for critically ill patients in ICU. The conventional clinical practice has been over reliant on Hb [1–3], where an increase in post-transfusion Hb is oblivious in anemic patients. Additionally, from Tables 1 and 2, we observed a decreasing effect in baseline Hb and hematocrit ranges as we move from A to D population. Where patients in group D with highest baseline SF are least critical, characterized by smallest Sequential Organ Failure Assessment (SOFA) max total score, and patients in A with lowest baseline SF are most critical, reflecting large SOFA scores. Thus, an inability to uncover the impact on respiratory physiology subsequent to RBC transfusion makes Hb incapable to show the transfusion efficacy for ICU patients. To strengthen Hb-reliant conventional approach, our study introduced SF as a surrogate and showed that effectiveness of the RBC transfusion is strongly associated with the pre-transfusion SF ratio. However, future prospective studies and clinical trials may be required to validate the hypothesis for its more generalizability.

Based on this initial study, we demonstrate that pre-transfusion SF ratio is significantly associated with RBC transfusion response, introducing SF ratio as a measure for RBC transfusion efficacy. Additionally, we show that SF ratio is not meaningfully correlated to SpO_2_ or Hb. Our results suggest that additional studies evaluating the SF ratio as a biomarker to determine the response to RBC transfusion (e.g. ΔSF) and the neediest RBC transfusion recipients (e.g. SF<100) are warranted. Our findings are consistent with studies showing the lack of improvement in oxygenation following RBC for many critically ill patients and emphasize the need for pragmatic, readily available measures to more precisely identify those critically ill patients who are most likely to benefit from RBC transfusion.

## Data Availability

All data produced in the present study are available upon reasonable request to the authors.

## Acknowledgements

The authors would like to thank the anonymous reviewers in advance for their useful suggestions and comments. R Kamaleswaran and T Choudhary were supported by the National Institutes of Health under Award Numbers R01GM139967, R21GM151703, and R21GM14893.

## Financial disclosures and conflicts of interest

R Kamaleswaran and T Choudhary were supported by the National Institutes of Health under Award Numbers R01GM139967, R21GM151703, and R21GM14893. The authors declare no competing financial interests.

## Authors contributions

R.K. conceived of the study and developed the study design. T.C. performed data and research analysis and wrote the manuscript; R.M.P. and R.K. helped with interpretation, editing of the manuscript, and read and approved the final manuscript; C.D.J., G.S., and J.D.R. read, suggested feedbacks, and approved the final manuscript.

## Notes

### Competing Interest Statement

The authors have declared no competing interest.

### Author Declarations

IRB of Emory University gave ethical approval for this work.

