## Supplemental File for "SpO_2_/FiO_2_ Ratio as a Better Metric for Assessment of RBC Transfusion Effectiveness in Non-traumatic Critically Ill Patients with Physiologic Derangements"

**Short title:** SF Ratio as a marker to evaluate RBC transfusion efficacy in critical illness

**E-mail addresses:** (Tilendra Choudhary), (Geoffrey Smith), (John D. Roback), (Ravi M. Patel), (Cassandra D. Josephson), (Rishikesan Kamaleswaran)

**Table of Contents**

1. eFigure 1 ……………………………………………………………………………….….3
2. eFigure 2. …………………………………………………………………….……………4

This supplementary document includes two figures with reference to the manuscript. eFigure 1 illustrates a localized study on Emory patients having different ranges of baseline SF ratios. After the transfusion event, a change in SF is measured and presented with baseline hemoglobin for three post-transfusion segments T1, T2 and T3. We observed that patients with the lowest baseline SF ratio (1-100), reflecting those most critically ill, consistently demonstrated an improvement in SF ratio subsequent to RBC transfusion. For patients with a baseline SF >300, reflecting the population with the least respiratory compromise, no significant improvement in SF ratio was observed following RBC transfusion, and in fact respiratory physiology worsened after transfusion. eFigure 2 illustrates the same study on Grady patients’ data for the validation purpose.

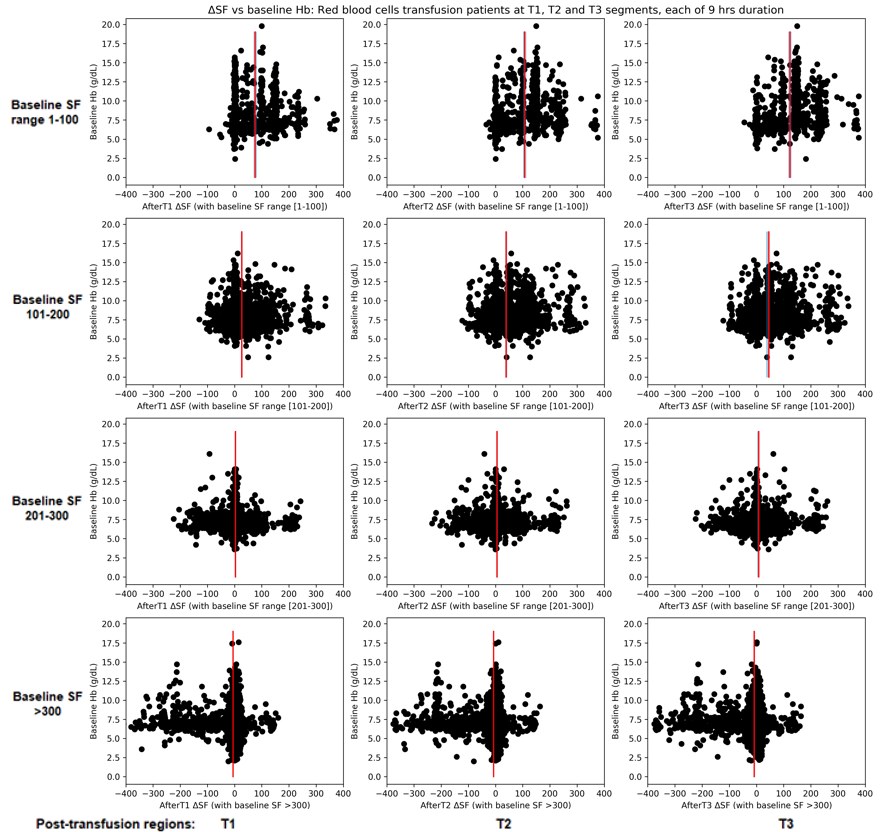

**eFigure 1:** **Emory data:** **Localized study on patients having different ranges of baseline SF ratios.** ΔSF vs baseline Hb for three consecutive post-transfusion regions (each of duration 9 hrs) with baseline SF range [row-1] 1-100, [row-2] 101-200, [row-3] 201-300, and [row-4] >300. Note that data-points denote the distribution of patients. Red vertical line with blueish filled region around it represents the mean difference in SF with 95% confidence interval (CI).

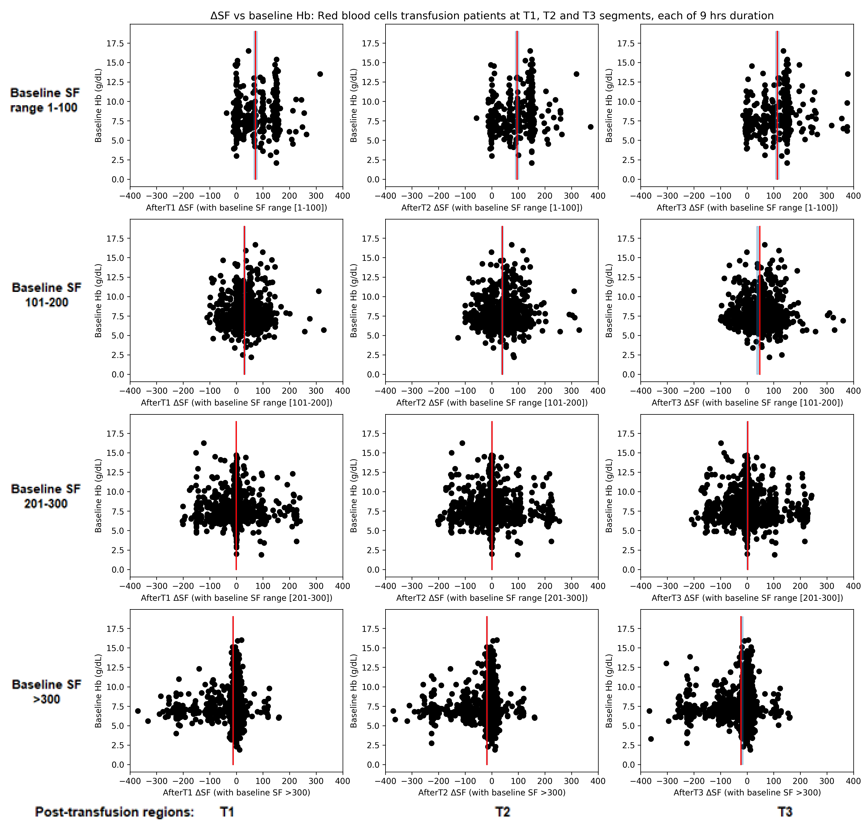

**eFigure 2:** **Grady data:** **Localized study on patients having different ranges of baseline SF ratios.** ΔSF vs baseline Hb for three consecutive post-transfusion regions (each of duration 9 hrs) with baseline SF range [row-1] 1-100, [row-2] 101-200, [row-3] 201-300, and [row-4] >300. Note that data-points denote the distribution of patients. Red vertical line with blueish filled region around it represents the mean difference in SF with 95% confidence interval (CI).
